# Epigenetic predictors of lifestyle traits applied to the blood and brain

**DOI:** 10.1101/2020.11.27.20239764

**Authors:** Danni A. Gadd, Anna J. Stevenson, Robert F. Hillary, Daniel L. McCartney, Nicola Wrobel, Sarah McCafferty, Lee Murphy, Tom C. Russ, Sarah E. Harris, Paul Redmond, Adele M. Taylor, Colin Smith, Jamie Rose, Tracey Millar, Tara L. Spires-Jones, Simon R. Cox, Riccardo E. Marioni

**Author notes:** **Correspondence:** Riccardo Marioni, Centre for Genomic and Experimental Medicine, Institute of Genetics and Molecular Medicine, University of Edinburgh, Edinburgh, UK.

## Abstract

Modifiable lifestyle factors influence the risk of developing many neurological diseases. These factors have been extensively linked with blood-based genome-wide DNA methylation (DNAm), but it is unclear if the signatures from blood translate to the target tissue of interest - the brain. To investigate this, we apply blood-derived epigenetic predictors of four lifestyle traits to genome-wide DNAm from five post-mortem brain regions and the last blood sample prior to death in 14 individuals in the Lothian Birth Cohort 1936 (LBC1936). Using these matched samples, we found that correlations between blood and brain DNAm scores for smoking, high density lipoprotein (HDL) cholesterol, alcohol and body mass index (BMI) were highly variable across brain regions. Smoking scores in the dorsolateral prefrontal cortex had the strongest correlations with smoking scores in blood (r=0.5, n=14) and smoking behaviour (r=0.56, n=9). This was also the brain region which exhibited the strongest correlations for DNAm at site cg05575921 - the single strongest correlate of smoking in blood - in relation to blood (r=0.61, n=14) and smoking behaviour (r=-0.65, n=9). This suggested a particular vulnerability to smoking-related differential methylation in this region. Our work contributes to understanding how lifestyle factors affect the brain and suggests that lifestyle-related DNAm is likely to be both brain region dependent and in many cases poorly proxied for by blood. Though these pilot data provide a rarely-available opportunity for the comparison of methylation patterns across multiple brain regions and the blood, due to the limited sample size available our results must be considered as preliminary and should therefore be used as a basis for further investigation.

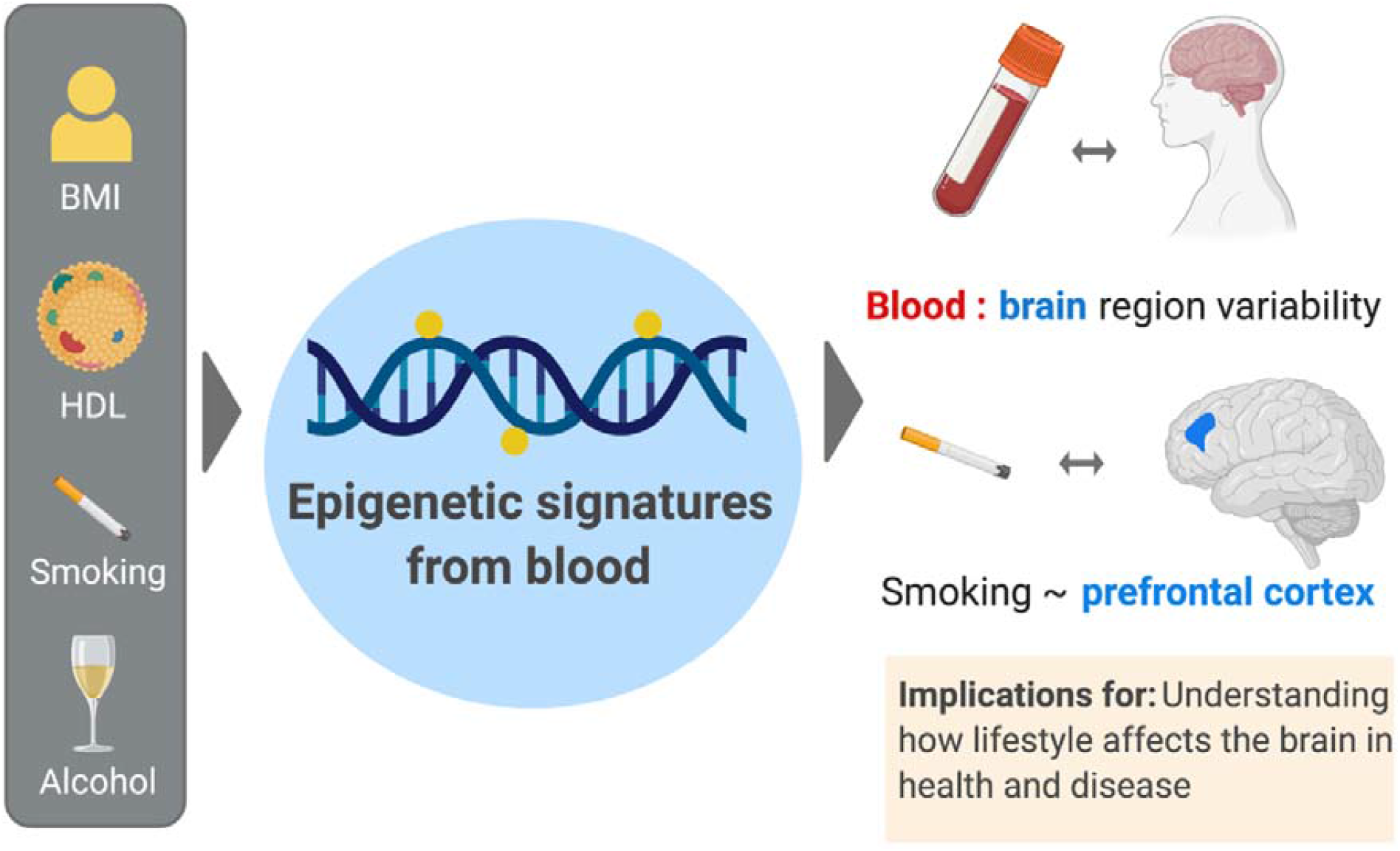

**Graphical abstract 203mm x 127mm (DPI 300)**

**Abbreviated summary [50 words]: We apply blood-derived epigenetic signatures of lifestyle traits to matched blood and brain samples, uncovering variability in how well blood translates across brain regions and a relationship between smoking and the prefrontal cortex**. Our preliminary results contribute to understanding how lifestyle-related DNA methylation affects the brain in health and disease.

## 1 Introduction

DNA methylation (DNAm) is one route by which modifications to the genome can occur and typically involves the addition of a methyl group to a cytosine residue on CG dinucleotides (CpGs) (Mazzio and Soliman, 2012). Lifestyle traits such as smoking, alcohol intake and body mass index (BMI), as well as High density Lipoprotein (HDL) cholesterol levels are known to associate with differential blood DNAm at CpG sites across the genome (Joehanes *et al*., 2016; Sayols-Baixeras *et al*., 2016; Braun *et al*., 2017; Mendelson *et al*., 2017; Sundar *et al*., 2017; Wahl *et al*., 2017; Liu *et al*., 2018). These lifestyle factors are associated with a range of brain health outcomes and neurological diseases (Donnan *et al*., 1993; Kivipelto and Solomon, 2006; Helzner *et al*., 2009; Bove *et al*., 2016; Quek *et al*., 2017; Sabia *et al*., 2018), in addition to brain morphology differences (Wobrock *et al*., 2009; Karama *et al*., 2015; Opel *et al*., 2015; Corley *et al*., 2019). Whereas the DNAm differences are likely to be a consequence, as opposed to cause, of lifestyle traits, it is unknown if these patterns are consistent across the blood and brain. Previous work suggests that blood is unlikely to reflect brain reliably for all CpGs, but that some sites more closely reflect brain DNAm than others (Hannon *et al*., 2015; Edgar *et al*., 2017; Braun *et al*., 2019). Given that the brain is the critical organ of interest for the pathology of neurological diseases, characterising the signature of DNAm resulting from lifestyle exposures in brain, as well as the extent to which blood DNAm can proxy for this is therefore paramount.

Blood-based DNAm predictors have been previously shown to explain 60% of the variance in self-reported smoking patterns and ∼12% of the variance in the alcohol, smoking, and BMI when projected into blood DNAm (McCartney *et al*., 2018). We therefore hypothesised that differential methylation patterns at the CpG sites associated with lifestyle traits in these blood predictors would be present across the corresponding sites in the brain. To test this, we applied the described out-of-sample blood-based predictors from our previous work to a pilot dataset consisting of matched blood and brain samples in 14 individuals from the Lothian Birth Cohort 1936 (LBC1936). We profiled four traits: HDL, BMI, alcohol and smoking. The most recent DNAm measure in blood taken prior to death was matched with genome-wide DNAm from *post-mortem* brain samples across five regions: Brodmann’s areas BA35 (hippocampus), BA46 (dorsolateral prefrontal cortex), BA24 (anterior cingulate cortex), BA20/21 (inferior temporal cortex) and BA17 (primary visual cortex).

We first assessed the correspondence between the scores derived from blood-based lifestyle predictors between the blood and brain, to characterise how well circulating DNAm measures of lifestyle traits may translate to the brain. We then compared the associations between the predictor scores and the corresponding self-reported or clinically assessed data available for each lifestyle trait. DNAm at a single CpG (cg05575921 in the *AHRR* gene; the strongest CpG correlate of smoking in blood which is hypomethylated in response to smoking (Fasanelli *et al*., 2015; Bojesen *et al*., 2017)) was also profiled in each of these analyses. This site is a key component of the smoking predictor and we hypothesised that the same hypomethylation observed in blood would also be evident in the brain. The relationship between blood-based predictor scores and lifestyle traits are presented for 499 individuals in the wider LBC1936 group to illustrate how representative the 14 individuals are of a larger reference group. The brain regions and study design are presented in **Fig. 1**.

**Fig. 1.**
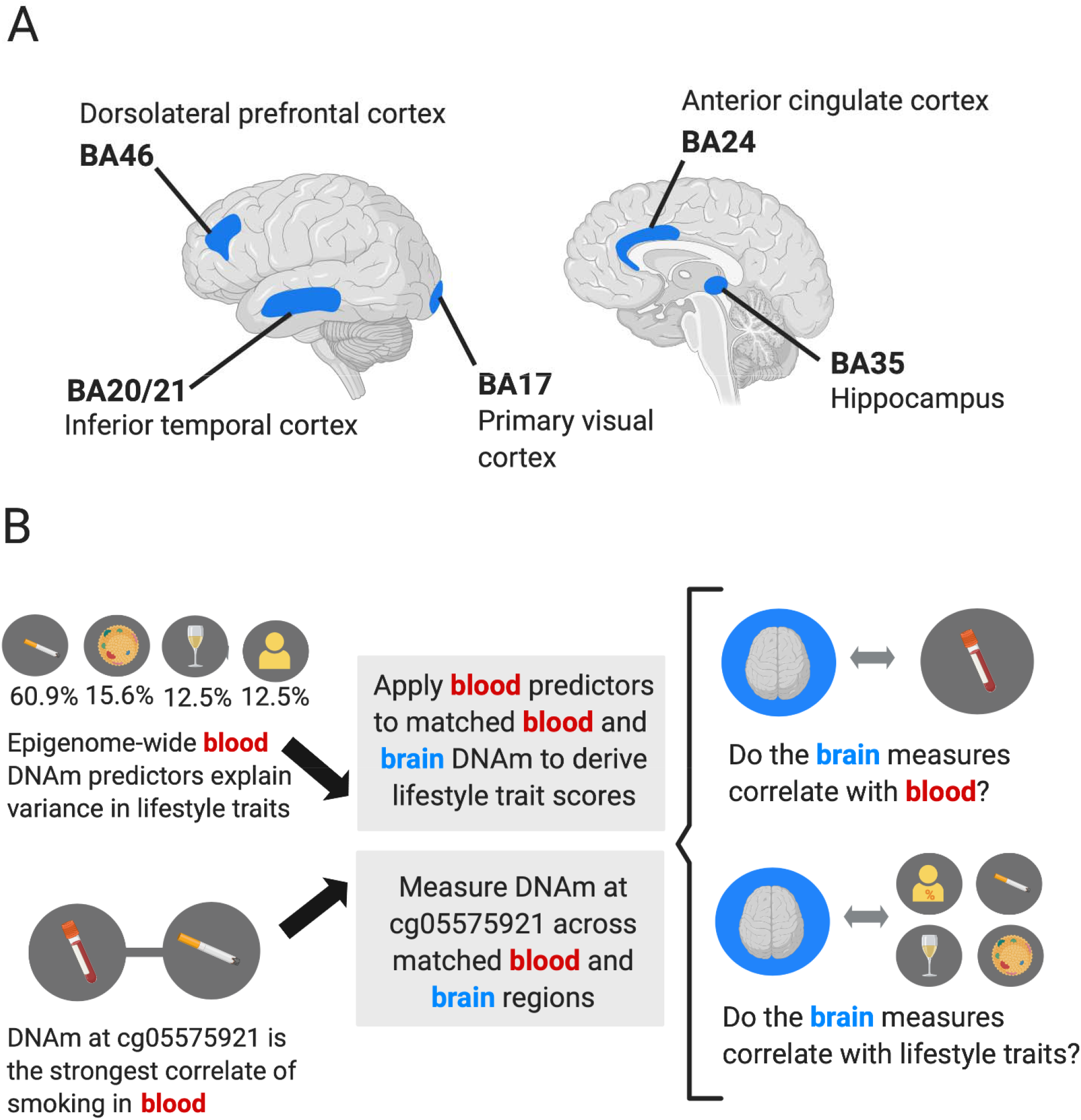
The application of blood-derived epigenetic predictors of four lifestyle traits to matched blood and brain DNA methylation samples in 14 individuals. (**A)** The five brain regions examined in 14 individuals with matched DNAm across brain regions and the last blood sample taken prior to death. (**B)** Epigenetic predictors of HDL, BMI, alcohol and smoking generated in 5087 out-of-sample individuals (McCartney *et al*., 2018) were applied to matched genome-wide blood and brain DNAm. The cg05575921 site in the *AHRR* gene locus was also measured across matched blood and brain samples. Analyses investigated the correlation between blood and brain measures and the correlation between the measures and respective lifestyle trait phenotypes relevant in each case. Figure created with BioRender.com.

## 2 Methods

### 2.1 The Lothian Birth Cohort 1936

The Lothian Birth Cohort 1936 (LBC1936, N=1,091) is a longitudinal study of ageing individuals who reside in Edinburgh and the surrounding areas (Deary *et al*., 2012; Taylor *et al*., 2018). Participants completed a childhood intelligence test at age 11 years in 1947 and were recruited for this cohort at the mean age of 70 years. They have been followed up approximately every 3-4 years since (currently at the 6^th^ wave), collecting a series of cognitive, clinical, physical and social data, along with blood donations that have been used for genetic, epigenetic, and proteomic measurement. Approximately 15% of individuals in the LBC1936 have consented to *post-mortem* tissue collection. To date, the samples from 14 individuals are available and were therefore selected as the brain bank group (n=14) for the present study.

### 2.2 Ethics

Written informed consent was obtained from all participants. Ethical permission for the LBC1936 was obtained from the Multi-Centre for Scotland (MREC/01/0/56), Lothian (LREC/2003/2/29) and Scotland A (07/MRE00/58) Research Ethics Committees. Use of human tissue for post-mortem studies has been reviewed and approved by the Edinburgh Brain Bank ethics committee and the ACCORD medical research ethics committee, AMREC (ACCORD is the Academic and Clinical Central Office for Research and Development, a joint office of the University of Edinburgh and NHS Lothian, ethical approval number 15-HV-016). Human tissue from the Edinburgh brain bank was used under the research ethics committee (REC) approval (16/ES/0084. All experimental methods were in accordance with the Helsinki declaration.

### 2.3 Blood DNAm in the Lothian Birth Cohort 1936

DNA from whole blood at 485,512 CpG sites was assessed using the Illumina Human Methylation 450K array at the Edinburgh Clinical Research Facility. The full details of the processing steps have been previously described (Shah *et al*., 2014; Zhang *et al*., 2018). Raw intensity data were background-corrected and normalised using internal controls. Following background correction, manual inspection permitted removal of low quality samples presenting issues relating to bisulphite conversion, staining signal, inadequate hybridisation or nucleotide extension. Quality control analyses were performed to remove probes with low detection rate <95% at P<0.01. Samples with a low call rate (samples with <450,000 probes detected at p-values of less than 0.01) were also eliminated. Furthermore, samples were removed if they had a poor match between genotype and SNP control probes, or incorrect DNA methylation-predicted sex. There were a total of 450,276 probes which remained.

Blood DNAm was available for up to 4 waves, measured over a 10-year period. The most recent blood sample prior to death was selected and self-report and clinical information was also taken from the most recent wave for which it was available. The most recent blood measurement was performed at wave 3 in seven individuals and wave 4 in the remaining seven, with a mean time between blood sampling and death of 3.2 years (SD 1.6) in the wave 3 group and 1.6 years (SD 0.94) in the wave 4 group (**Supplementary Table 1**). In the 14 individuals, the mean age at blood sampling was 77.9 (SD 1.7) and the mean age at death was 80.3 (SD 1.6). The blood DNAm reference group (n=499) was taken from wave 4 of the LBC1936 and the mean age at sampling was 79.3 years (SD 0.62). Lifestyle trait information and blood DNAm were recorded in a consistent way across both the brain bank and reference groups.

### 2.4 Brain DNAm processing in the Lothian Birth Cohort 1936

Brain tissue samples were received from the Edinburgh Brain Bank. Regions of the brain were dissected after brains were removed and cut coronally into slices, as per previous methodology (Samarasekera *et al*., 2013). Samples from regions BA46 (dorsolateral prefrontal cortex), BA17 (primary visual cortex), BA24 (anterior cingulate cortex), BA20-21 (ventral (20) and lateral (21) inferior temporal cortex) and BA35 (hippocampus) were taken from the cortex and snap frozen. A tissue selection of ∼25mg was processed for DNA extraction, which was done using the DNeasy kit (Qiagen). DNAm was measured at the Edinburgh Clinical Research Facility using Illumina MethylationEPIC BeadChips. Quality control steps were then performed: the wateRmelon pfilter() function removed samples in which >1% of probes had a detection p-value of >0.05, probes with a beadcount of <3 in >5% of samples, and probes with >1% of samples reaching a detection p-value of >0.05. Additional SNP probes and cross-hybridising probes on X and Y chromosomes were removed (McCartney *et al*., 2016). If a discordance between the methylation-predicted sex and recorded sex was identified samples were also removed. Performance of 15 normalisation functions was examined, as per Pidsley et al., with danet selected as the top-ranking method (Pidsley *et al*., 2013).. The normalised data had a total of 69 samples (14 individuals, across 5 regions, with 1 hippocampal sample unavailable) and 807,163 probes. DNAm beta values were used in all analyses.

### 2.5 Lifestyle trait information in the Lothian Birth Cohort 1936

Lifestyle trait phenotype measurements were as follows: self-reported smoking status (0=never smoked, 1=former smoker, 2=current smoker); alcohol consumption in a usual week (converted into units); BMI (defined as the ratio of weight in kg divided by height in m^2^) and HDL cholesterol (measured in mmol/L). Pack years smoked was calculated by multiplying the number of packs of cigarettes smoked per day by the number of years the individual had smoked for, divided by 20 (cigarettes per pack). If a person reported having never smoked, their pack years was recorded as 0 and this was done for both the reference and brain bank groups. Of the reference group, 37 people had pack years available, 193 did not have information available and the remaining 270 indicated that they were never smokers. Five individuals were excluded from the pack years trait in the brain bank group as they did not have information on the number of packs smoked per day, which is why a smaller subset were used for analysis of this trait (n=9). One individual in the brain bank group did not have a starting age for smoking; however, the mean of the group starting age was imputed (16 years old). Any other unknown trait information from the reference group was not used. Mortality data were obtained through data linkage to the National Health Service Central Register, provided by the General Register Office for Scotland (now National Records of Scotland) and were correct as of July 2020.

### 2.6 DNAm signature predictor scores for lifestyle traits

CpG predictor weights for complex traits were generated and validated through a pipeline described previously (McCartney *et al*., 2018). Briefly, LASSO penalised regression was used to identify a linear combination of CpG sites that associated with lifestyle traits. The beta coefficients generated for the CpG sites, known as weights, were based on whole-blood derived samples from 5087 individuals from the Generation Scotland cohort (Habota *et al*., 2019). Here, the described CpG weights (taken from McCartney et al, 2018. Additional file 1: Tables S1-3 and Table S6) were applied to DNAm at the same CpG sites in LBC1936 individuals and summed to generate predictor scores for smoking, HDL, BMI and alcohol. This was done for DNAm in the blood and five brain regions in the brain bank group (n=14) and the blood DNAm which was available in the reference group (n=499).

### 2.7 Statistical analyses

First, Pearson correlations were applied to measure the correspondence between the methylation predictors in the blood and brain. These correlated the lifestyle predictor scores - generated from the application of blood-derived CpG predictor weights - between blood and brain, as well as the DNAm measurements at site cg05575921 between the blood and brain. Second, Pearson correlations were used to measure the relationships between brain and blood DNAm measures of lifestyle traits and the respective lifestyle trait information. Units of alcohol consumed weekly, BMI, HDL cholesterol from blood and both smoking status and pack years smoked were included as traits and correlated with the DNAm-based lifestyle predictor scores. Smoking information was also correlated with DNAm at site cg05575921. These lifestyle trait correlations were also performed for the blood-based predictor scores and cg05575921 measurements in the reference group from wave 4 of the LBC1936.

As a sensitivity analysis, Spearman correlations were conducted for every association tested in the described Pearson correlations. Inter-region Spearman correlations were also performed for the blood-derived lifestyle predictor scores applied to brain and for the cg05575921 DNAm measure across the five regions. Though covariate information was available on the proportion of neurons in the brain, the brain weight, brain pH and most-mortem interval, power calculations suggested that linear mixed effects regression analyses in the sample size of 14 individuals were unlikely to be sufficiently powered to detect significant effects **(Supplementary Table 2)**. For this reason, no further statistical testing was performed. Correlations were conducted using the Hmisc library (Version 4.4-0) (Harrell, 2020). Inter-region correlation heatmaps were produced using the ggcorrplot library (Version 0.1.3) (Kassambara, 2019) and correlation plots were produced using the corrplot library (Version 0.84) (Wei and Simko, 2017). All analyses were performed using R (Version 3.6.3) (R-Core-Team and RCore-Team, 2015).

### 2.8 Data availability

LBC1936 data are available on request from the Lothian Birth Cohort Study, University of Edinburgh. LBC1936 data are not publicly available due to them containing information that could compromise participant consent and confidentiality.

## 3 Results

### 3.1 Cohort assessment

Summary information for the 14 individuals from the LBC1936 brain bank subset and the reference group (n=499) is presented in **Supplementary Table 3**. The brain bank subset had a higher proportion of males (64%) than the reference group (50%). Age at death in the brain donor group ranged from 77.6 to 83.0 years and the mean age of the reference group was well matched to this group (77.90 vs 79.31, respectively). The majority of the 14 individuals within the brain bank subset either had been or were still smokers (86%) at the time of death, which was a higher proportion than in the reference group (46%). Most of the 14 individuals (86%) had high HDL cholesterol (> □ 1 mmol/L), drank alcohol weekly (92%) and had mean BMI of 25.5 kg/m^2^. Five of the individuals did not have smoking pack years data recorded.

### 3.2 Correlation between blood and brain measures

Blood-derived epigenetic predictors for lifestyle traits were applied to the matched blood and brain DNAm samples in the brain bank group to generate lifestyle trait scores. DNAm at the smoking-associated CpG site cg05575921 was also considered. The variability across brain regions in the magnitude of correlations between DNAm predictor scores and DNAm at site cg05575921 are illustrated in **Supplementary Fig. 1**. The correlations between both the blood and brain lifestyle scores and the blood and brain measures of cg05575921 were regionally variable (**Fig. 2**). The strongest association between blood and brain DNAm at site cg05575921 was observed for BA46 (r=0.61, n=14) followed by BA17 (r=0.39, n=14). Blood smoking scores were most highly correlated with the BA46 region scores for smoking (r=0.5, n=14), with weaker associations observed for BA24 (r=0.29, n=14) and BA35 (r=0.36, n=13). BMI scores were negatively correlated in regions BA46 (r=-0.72, n=14) and BA35 (r=-46, n=14), suggesting that methylation patterns related to BMI were divergent across blood and brain. The HDL scores were moderately-correlated between the blood and region BA20/21 (r=0.55, n=14), with weaker correlations observed for regions BA24 (r=0.38, n=14) and BA35 (r=-0.32, n=13). Blood alcohol scores were most highly correlated with BA24 (r=0.35, n=14) and in BA46 (r=0.25, n=14) alcohol scores. A sensitivity analysis suggested that the strongest correlations were consistent across Pearson and Spearman methods. The correlation coefficients and p-values are presented for both analyses in **Supplementary Table 4**.

**Fig. 2.**
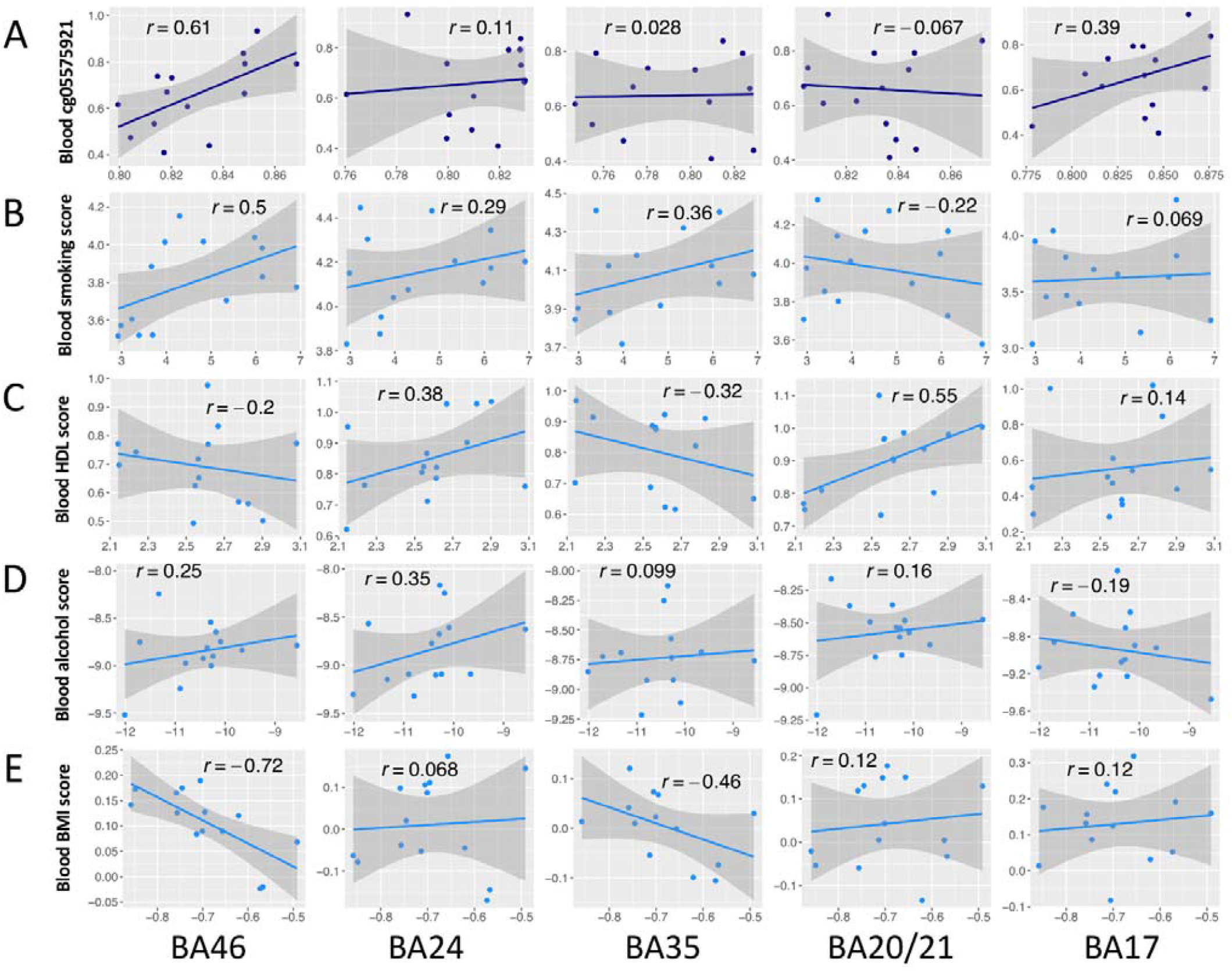
Correlations for (A) site cg05575921 and blood-derived lifestyle trait predictor scores for (B) smoking, (C) HDL, (D) alcohol and (E) BMI traits applied to the blood and brain. Relationships between brain DNAm and blood DNAm are shown for each brain region and measure. Each point represents one individual. Pearson correlation coefficients are annotated in each case. All individuals had both blood and brain samples available (n=14), apart from one individual for which no BA35 hippocampal sample was available (n=13). The solid blue line represents the linear regression slope; shaded areas represent 95% confidence intervals.

### 3.3 Correlation of blood and brain measures with lifestyle traits

The lifestyle trait scores generated by applying blood-derived epigenetic predictors to brain and blood samples and the DNAm measures at site cg05575921 were then correlated with clinical and self-reported lifestyle phenotypes (**Fig. 3**).

**Fig. 3.**
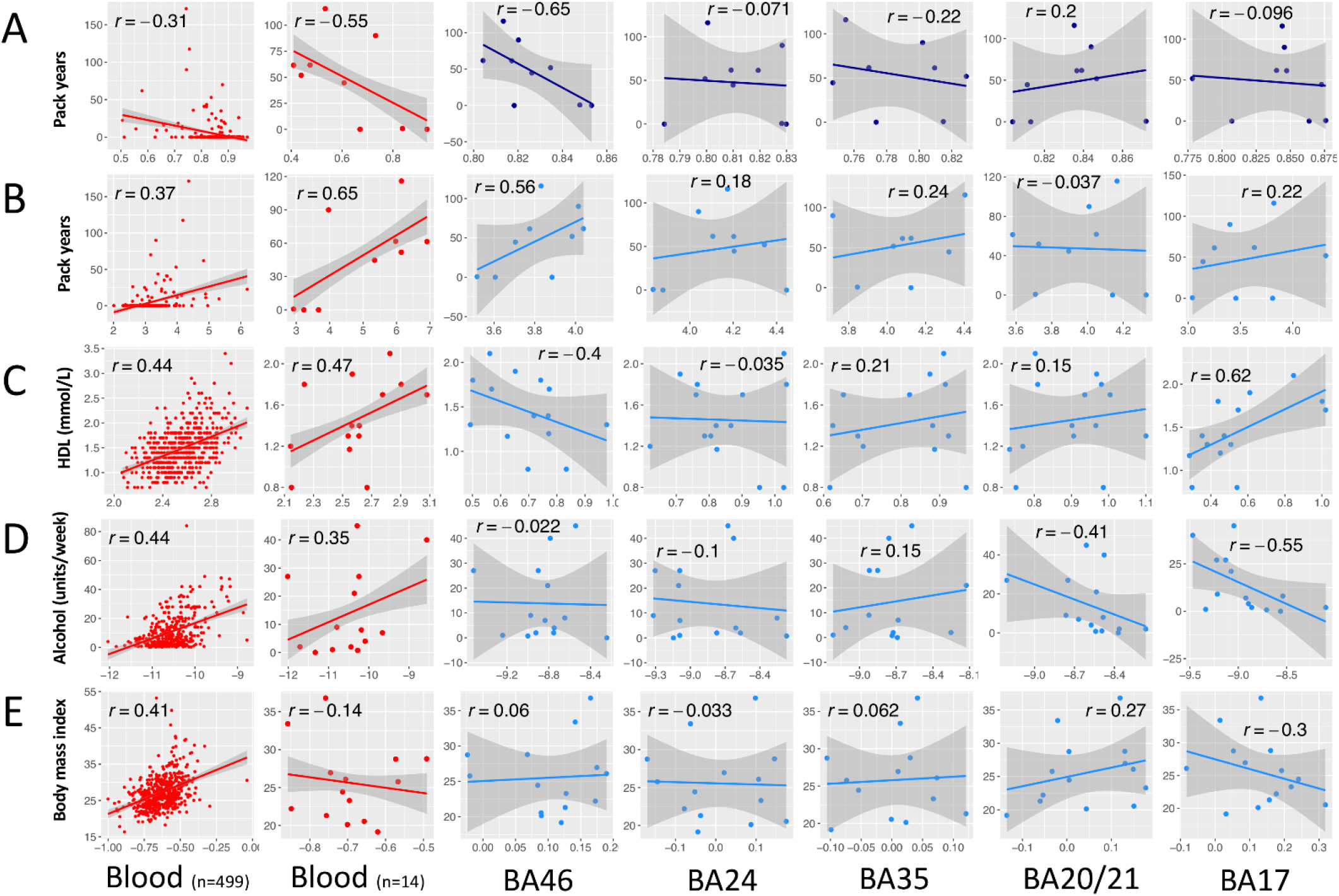
Lifestyle trait correlations with the blood DNAm measures from the reference group (up to n=499) and the blood and brain measures for the brain bank group (n=14). Correlations for blood measures are shown in red, with cg05575921 DNAm in dark blue and lifestyle trait scores generated from the application of blood-derived lifestyle predictors to brain in light blue. **A**, DNAm at site cg05575921 is correlated with pack years smoked for the reference (n=306) and brain bank (n=9) groups. Correlations are then provided for each group between lifestyle trait scores for (**B**) smoking, (**C**) HDL, (**D**) alcohol and (**E**) BMI, against relevant lifestyle trait information. HDL is measured in mmol/L. Alcohol is measured in average units per week. Each point represents one individual. Pearson correlation coefficients are annotated in each case. All individuals had both blood and brain samples available (n=14), apart from one individual for which no BA35 hippocampal sample was available. The solid blue line represents the linear regression slope; shaded areas represent 95% confidence intervals.

As expected, in the reference cohort (n=499) we observed hypomethylation in blood DNAm levels of cg05575921 associated with pack years smoked for current and former smokers (r=-0.31, n=307, p < 0.05). This trend was mirrored by the negative association found for blood DNAm at cg05575921 in the brain bank subset (r=-0.55, n=9). Similar patterns were observed in some but not all brain regions. The strongest association for cg05575921 in brain with pack years was found for region BA46 (r=-0.65, n=9), followed by a weaker correlation in BA35 (r=-0.22, n=8). BA20/21 had an opposite trend to that expected in blood (r=0.2, n=9). Methylation in all other brain regions showed much weaker associations with the pack years phenotype (|r| ≤ 0.1).

Regarding the lifestyle predictor scores, all trait scores in the blood samples from the brain bank subset were reflective of the wider reference cohort in terms of direction and approximate magnitude (|r| between 0.55 and 0.65), except BMI for which the brain bank group (r=-0.14, n=14) was not representative of the reference group (r=0.41, n=499). As with the results for cg05575921, BA46 was the region for which smoking scores were most highly correlated with pack years smoked (r=0.56, n=9). To a lesser extent BA17 (r=0.22, n=9) and BA35 (r=0.24, n=8) smoking scores were also correlated with pack years. HDL brain scores correlated with HDL trait information strongly in BA17 (r=0.62, n=14), whereas BA35 (r=0.21, n=14) had a weaker association and a negative trend was observed for BA46 (r=-0.4, n=14). Alcohol signatures showed the strongest trends in regions BA17 (r=-0.55, n=14) and BA20/21 (r=-0.41, n=14), in the opposite direction to that found in blood. BMI signature scores in BA17 were negatively correlated with BMI information (r=-0.3, n=14) in a trend opposite to the LBC reference group and BA20/21 also had a positive correlation with BMI (r=0.27, n=14). The remaining regions did not show any notable correlations with BMI trait information (|r| ≤ 0.1). A sensitivity analysis comparing Pearson and Spearman methods found that, though there was variability across the methods, the top associations were consistent (**Supplementary Table 5**). Correlations between smoking measures and smoking status are also available in this file.

## 4 Discussion

Through the application of blood-derived epigenetic predictors of four lifestyle traits to whole-genome DNA methylation in matched blood and brain samples, we uncover regional variability in how well blood-derived scores may be able to proxy for brain-based scores. Though our results highlight disparities between the blood and the brain, we did find evidence to suggest that the blood-derived predictors of lifestyle traits may translate to specific regions. The dorsolateral prefrontal cortex (BA46) was identified as a region of interest for the smoking trait and showed relationships in our analyses using the epigenetic predictor of smoking and the CpG site cg05575921 - the single strongest known correlate of smoking across the epigenome. We present these preliminary findings as a contribution to the ongoing, important question of how circulating DNAm measures are able to reflect methylation in brain tissue.

The pilot dataset used in this study provided a rarely-accessible resource to compare genome-wide methylation specific to each of the five brain regions against matched blood methylation in the same individuals. Given that the generation of epigenetic predictors requires large training datasets, it was not feasible to generate lifestyle predictors derived from brain DNA methylation in this study, but we hypothesised that CpG sites from the blood predictors of lifestyle traits would be concordant in their methylation patterns in the brain. We tested this by applying blood-derived CpG predictor weights to the matched blood and brain samples. This context is central to the interpretation of our results, which suggest that lifestyle-related methylation in some brain regions is more highly correlated with blood than others. The variability we found across brain regions may be due to several possibilities: 1) lifestyle traits may have a stronger influence on DNAm in the regions which were well-correlated, 2) poorly correlated regions may have CpGs influenced by lifestyle traits which are unique to the brain and therefore not captured through the blood predictors, or finally 3) poorly correlated regions may not have any distinct methylation patterns due to lifestyle traits. We believe that the latter is somewhat unlikely, as associations between lifestyle factors and both brain health outcomes and morphology differences have been documented across a range of studies (Donnan *et al*., 1993; Kivipelto and Solomon, 2006; Helzner *et al*., 2009; Wobrock *et al*., 2009; Karama *et al*., 2015; Opel *et al*., 2015; Bove *et al*., 2016; Quek *et al*., 2017; Sabia *et al*., 2018; Corley *et al*., 2019).

Previous studies have found that many sites in the genome are poorly correlated between the blood and brain with further variability across brain regions (Hannon *et al*., 2015; Edgar *et al*., 2017; Braun *et al*., 2019). The cg05575921 site has been shown to correlate between blood and adipose tissue (Tsai *et al*., 2018), suggesting that it may have a discernible trace across circulating measures and tissues; however, in most brain regions we did not observe strong correlations at this site both in relation to blood or smoking traits. Generally, there were weak correlations between the lifestyle predictors across blood and brain samples and in relation to lifestyle traits for many regions. Though the effects of lifestyle on brain DNAm directly are relatively unknown, in the LBC1936 cohort, the blood-based DNAm predictor for smoking used in the present study has been shown to associate with brain morphology differences (Corley *et al*., 2019). There is also evidence linking *in utero* exposure to smoking and alcohol use disorders to brain DNAm alterations (Chatterton *et al*., 2017; Gatta *et al*., 2019). Blood lipids have also been causally associated with DNAm differences in circulating cells (Dekkers *et al*., 2016), though it is unknown whether this is true of lipids and brain DNAm. Lipid measures from the blood have also been associated with brain volume and grey matter variability (Harris *et al*., 2020), though whether these trends are directly aligned to DNAm patterns is not characterised. Taken together, these studies suggest that our findings may be due to possibility (2), that there could be CpG sites related to lifestyle traits which are unique to the brain and may not be captured in the present study.

The dorsolateral prefrontal cortex (BA46) smoking predictor score and methylation at site cg05575921 were well-correlated with both the equivalent measures in blood and smoking trait information. A previous study found that of four regions, DNAm at cg05575921 in blood was most highly correlated with cg05575921 in the prefrontal cortex (r=0.28, P=0.02) in matched samples in 74 individuals (Hannon *et al*., 2015). Though this correlation is somewhat weaker than that observed in our study, it suggests that possibility (1) may be correct; there may be a particularly strong influence of smoking on differential DNAm in regions such as BA46. This possibility is further supported by studies that have pinpointed the frontal cortex as a region that is particularly vulnerable to the effects of smoking on brain morphology (Karama *et al*., 2015; Cox *et al*., 2019). One of the discussed studies was able to show that smoking had unique statistical contributions to brain morphology when modelling against many other vascular risk factors (VRFs) in a large population (N=9,722) (Cox *et al*., 2019); these findings indicate that there some brain areas may be more susceptible to smoking. As VRFs (such as smoking) associate with various adverse outcomes including cognitive ageing and dementia (Haley *et al*., 2018; Pasha *et al*., 2018; Sweeney *et al*., 2018; Cheng *et al*., 2020), areas such as BA46 may be more susceptible to ageing and effects that underpin cognitive function, as they show the strongest VRF-related coupling in blood and brain. A larger sample with more detailed regional sampling across the brain will be required to investigate this.

There are a number of limitations to our study. As discussed, the lack of brain DNAm samples meant that it was not possible to create lifestyle predictors in brain tissue. Recent work has generated predictors for ageing in cortical samples, providing evidence that, though imperfect, there is a concordance between predictors generated in blood and brain (Shireby *et al*., 2020). Future work should seek to determine if this is the case for lifestyle predictors. Second, there were only 14 individuals with matched samples in the LBC1936 brain bank. This meant that though we had information on covariates such as the *post-mortem* interval between death and brain DNAm sampling, regression analyses were not feasible due to insufficient power. Though confounders may have influenced our results, the effect of *post-mortem* interval is still debated (Ernst *et al*., 2008; Jarmasz *et al*., 2019). Third, the blood-based BMI DNAm signatures of the 14 individuals were not reflective of the wider reference group, which limits their interpretability. Fourth, many current or former smokers had not reported pack years information and there was variability in the strength of correlations across smoking phenotypes. Though trends for the smoking status trait were generally weaker than those observed for pack years, this difference may be reflective of the longitudinal nature of smoking that pack years captures. Fifth, whereas all participants were free from neurodegenerative conditions at the study recruitment at age 70, the absence of clinical data on subsequent clinical neurodegenerative diagnoses means that we cannot rule out the possibility that these results are partly driven by disease-specific DNAm profiles in the brain. Several studies have found differential DNAm at regions such as the hippocampus in those Alzheimer’s disease pathology, suggesting that DNAm alterations may result from the pathological changes seen before symptoms arise (De Jager *et al*., 2014)(Yu *et al*., 2015). Growing sample donations and ongoing clinical ascertainment will partly address these limitations in future work.

### Conclusion

In this study, we characterise variability in how well blood-derived epigenetic measures of lifestyle traits correlate when applied across a rarely-available pilot dataset consisting of matched blood and brain samples. We find variability in the alignment between blood and brain lifestyle predictor scores across brain regions, with the most notable relationships found between the dorsolateral prefrontal cortex (BA46) and smoking-related measures. Though our work relies on the application of blood-based signatures of lifestyle traits to brain tissue and is limited by low sample size, it nonetheless provides a preliminary insight into whether circulating DNAm proxies may reflect the epigenetic effects of lifestyle traits in the brain. This is critical given the known associations between modifiable lifestyle factors with both neurological disease risk and brain health outcomes.

## 5 Acknowledgements

The authors thank all LBC1936 study participants and research team members who have contributed, and continue to contribute, to ongoing LBC1936 studies.

## Abbreviations

BA17: Brodmann’s area 17
BA46: Brodmann’s area 46
BA24: Brodmann’s area 24
BA35: Brodmann’s area 35
BA20/21: Brodmann’s area 20/21
BMI: Body mass index
CpG: cytosine-guanine dinucleotide
DNAm: DNA methylation
HDL: High density lipoprotein
LBC1936: Lothian Birth Cohort 1936

