## Supplemental Figure 1 for "Epigenetic predictors of lifestyle traits applied to the blood and brain"

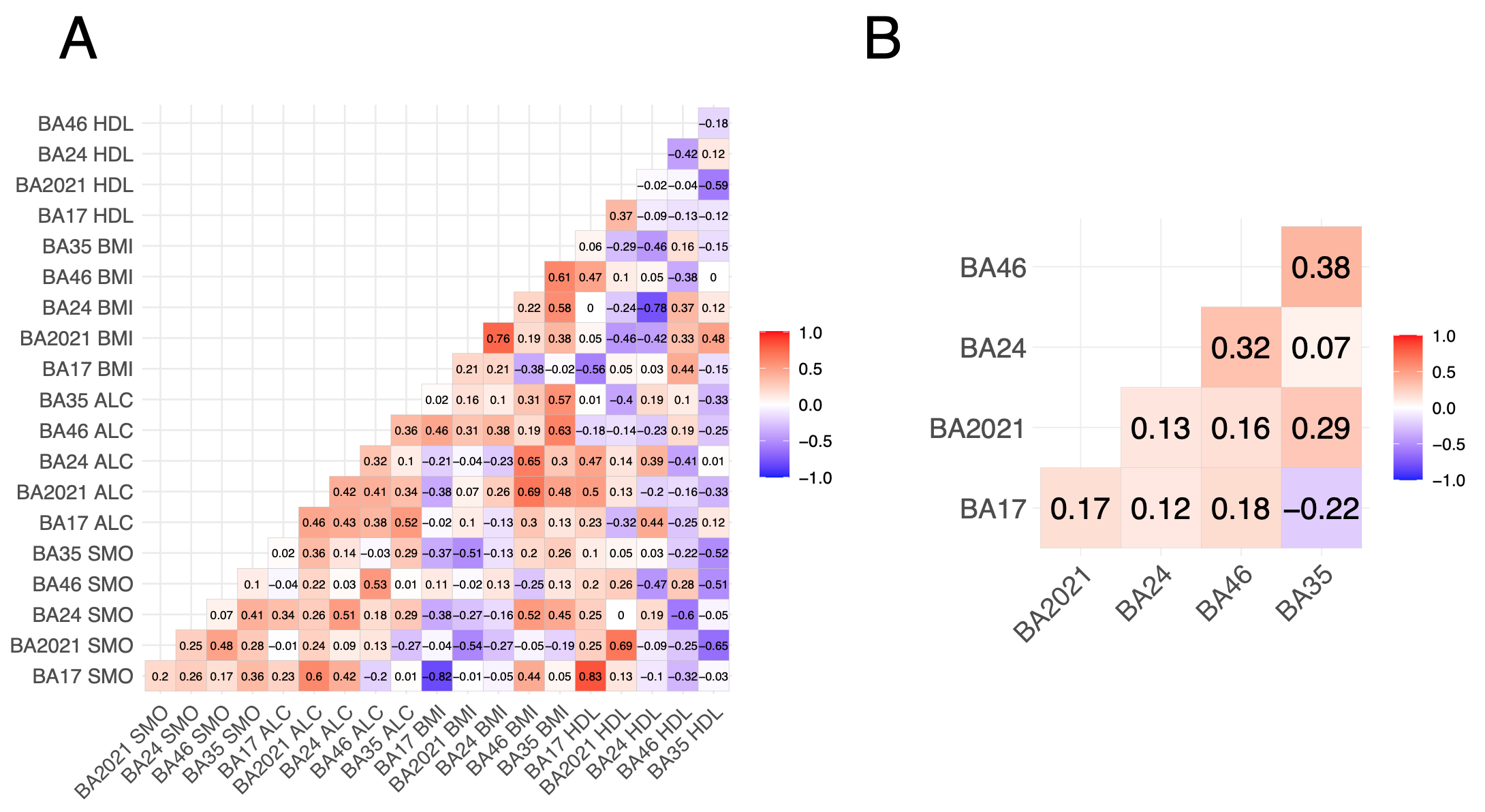
**Supplementary Fig. 1. Heatmaps showing correlations for (A) predictor scores and (B) DNA methylation at the CpG site cg05575921 across the five brain regions in the brain bank group.** Spearman correlation coefficients are presented in each case. There were 14 measurements available, except one individual for which hippocampal (BA35) sampling was not available. BA: Brodmann’s area. HDL: high density lipoprotein. BMI: body mass index. ALC: alcohol. SMO: smoking.
